# Trio exome sequencing as the stand-alone first-tier testing for prenatal diagnosis

**DOI:** 10.1101/2025.07.23.25331964

**Authors:** Haibo Li, Lulu Yan, Yuxin Zhang, Min Li, Jiangyang Xue, Liyun Tian, Juan Cao, Shanshan Wu, Yanling Dong, Yuying Zhu, Hongmei Zhou, Shuangshuang Shen, Xiayuan Xu, Guosong Shen, Xueping Shen, Penglong Chen, Yanting Xu, Shu Fang, Dandan Cai, Yuchao Wang, Danjun Liang, Juan Geng, Zhaojing Zheng, Changshui Chen

## Abstract

**BACKGROUND:** Whole exome sequencing (WES) is increasingly utilized in prenatal diagnosis of fetus with structural abnormality. However, evidence supporting its use as a first-tier genetic test for all pregnancies referred for invasive prenatal procedures remains limited.

**METHODS:** We conducted a multi-center prospective study enrolling 1,382 pregnancies from seven prenatal diagnosis centers all around China. Participants were divided into two groups based on clinical indications: those for whom CMA testing was traditionally recommended (CMA group) and those for whom CMA and WES were sequentially or concurrently recommended (CTW group) according to their indications. After excluding 200 pregnancies terminated prior to genetic test reporting, the cohort included 1,182 cases undergoing independent trio-WES and CMA testing. The primary outcome was determined through follow-up of at least 3 months after delivery.

**RESULTS:** Trio-WES yielded an overall diagnostic yield of 17.22%, with an incremental yield of 7.38% for SNVs/Indels not detected by CMA. Even for subjects in the CMA group, there’s a diagnostic yield of 4.02%, arising from SNVs/Indels that would be missed by CMA. Trio-WES showed a 100% concordance with CMA on detection of reportable cytogenomic events, with unique advantages over CMA in 6 cases with UPD and 3 cases with exon-level deletion. Importantly, WES results, whether positive, negative or uncertain, significantly impact clinical decision-making for pregnancy.

**CONCLUSIONS:** Our findings support the use of prenatal trio-WES as a stand-alone first-tier genetic test for all pregnancies referred for invasive prenatal diagnosis, regardless of indication.

## Introduction

Whole exome sequencing (WES) has emerged as a transformative diagnostic tool capable of detecting pathogenic variants beyond the scope of standard-of-care (SOC) genetic tests such as chromosomal microarray analysis (CMA)^1^, which remains the first-tier approach for invasive prenatal diagnosis in many healthcare systems constrained by conservative technology adoption frameworks. Although previous investigations have explored WES utility in selected prenatal cohorts—primarily restricted to fetuses with structural anomalies after negative CMA results^2, 3^—critical knowledge gaps persist regarding its performance across diverse clinical indications. Existing studies^4–6^ in low-risk pregnancies further limit generalizability by excluding cases with major CMA findings or established risk factors (e.g., biochemical screening abnormalities, advanced maternal age), thereby failing to represent the heterogeneous population seeking prenatal diagnosis. Crucially, while initial evidence suggests WES-based copy number variant (CNV) calling may approximate or even exceed microarray performance in pediatric settings^7^, rigorous validation of this methodology in prenatal contexts remains absent despite its potential to consolidate diagnostic workflows. This evidence deficit is particularly consequential given trio-WES’s unique cost-effective advantages in resource-limited environments, where integrated variant detection could alleviate burdensome multi-step testing protocols. Consequently, we conducted this multicenter prospective study, to our knowledge the first of its kind, to definitively establish the diagnostic yield, CNV concordance with CMA, and clinical utility of trio-WES as a stand-alone first-tier solution for unselected pregnancies undergoing invasive prenatal diagnosis.

## Methods

### STUDY DESIGN

This multicenter prospective study enrolled singleton pregnant women referred for invasive prenatal diagnosis due to maternal-fetal diverse risks across seven institutions (October 2022-April 2024, eFigure 1 in Supplement 1). Participants met ≥1 clinical criteria: (a) advanced maternal age, ≥35 years; (b) abnormal maternal serum biochemical screening; (c) abnormal non-invasive prenatal testing (NIPT); (d) abnormal ultrasound soft markers; (e) nuchal translucency (NT) ≥3.5mm; (f) ultrasound structural anomalies; (g) fetal growth retardation (FGR); (h) polyhydramnios/oligoamnios; (i) family history of genetic/likely genetic conditions; (j) adverse reproduction history; (k) exposure to teratogens; (l) other high risk conditions, e.g., pregnancies by in vitro fertilization (IVF) procedure. Pregnancy loss and stillbirth at enrollment were excluded from this study. Based on referral indications, subjects were stratified into two groups: 1) CMA group, in which CMA testing was traditionally recommended (standard indications: criteria a-d above), and 2) CTW group, in which CMA and WES were sequentially or concurrently recommended (expanded indications: criteria e-l). The primary objectives were to evaluate trio-WES as a first-tier diagnostic tool through assessment of: 1) overall diagnostic yield, 2) CNV detection concordance with CMA, and 3) clinical utility. The study was approved by the Ethics Committee of the seven participating centers (Supplement 1), written informed consent was obtained from every participant.

**Figure 1.**
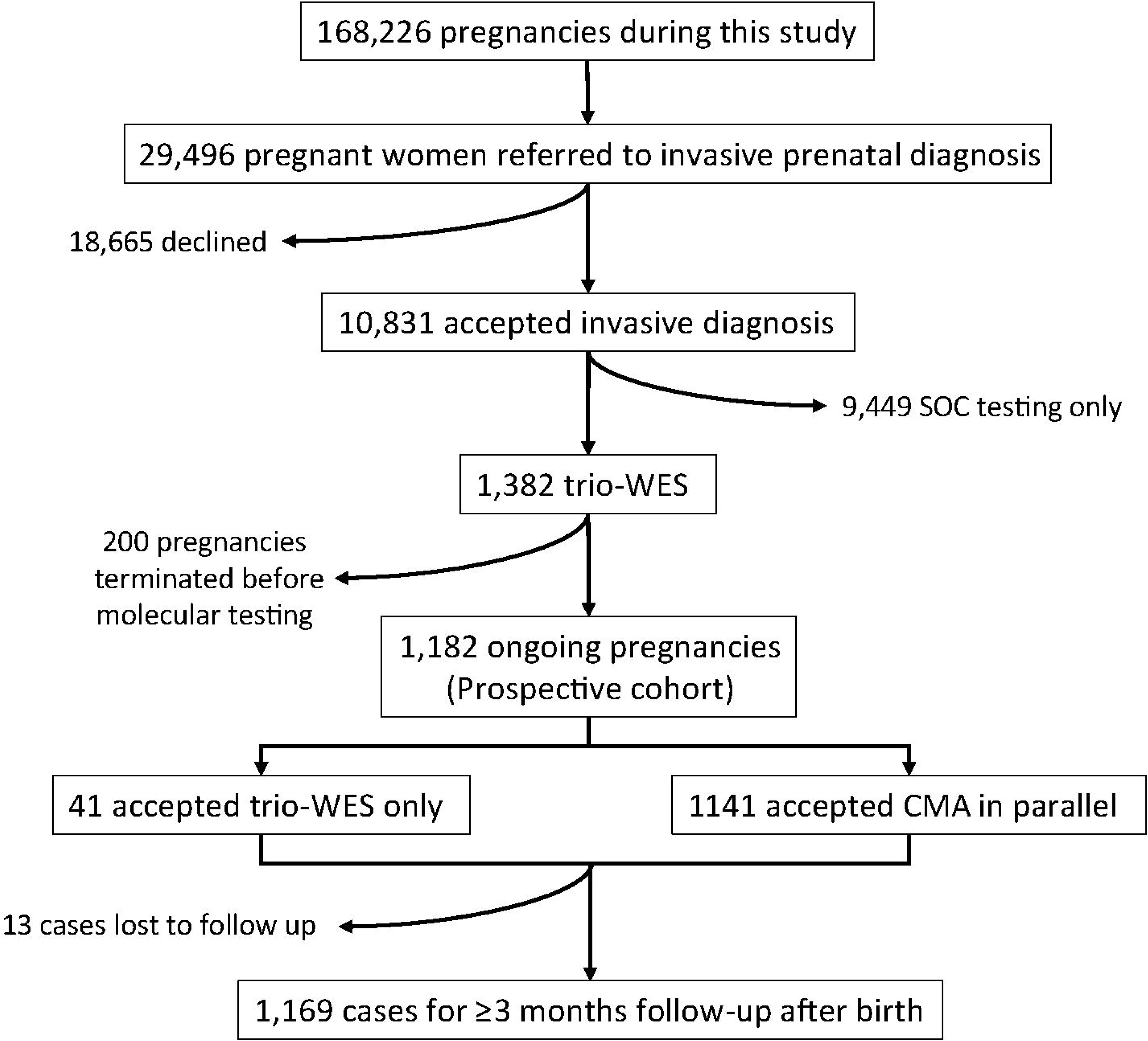
Study design and cohort description. According to the inclusion criteria of this study, a total of 1382 singleton pregnant women were enrolled from the seven prenatal diagnosis centers across China during October 2022 to April 2024 with a prospective study design. Amongst, 200 pregnancies were excluded from downstream analysis due to early termination before return of trio whole exome sequencing (trio-WES) testing results. A total of 1182 pregnancies constituted the prospective cohort, which was subjected to independent trio-WES and chromosomal microarray analysis (CMA) in parallel. The outcomes of the prospective cohort were determined through follow-up of at least 3 months postpartum. SOC: standard-of-care.

### EXOME SEQUENCING

Exome sequencing in this study was performed in Hangzhou Juno Genomics laboratories with a custom Twist Exome panel Omniseek^®^. Sequencing on DNBSEQ-T7 platforms achieved at least 100× mean coverage with 150-bp paired-end reads. Bioinformatic analysis included: 1) SNV/Indel calling via GATK-based pipeline, 2) CNV detection using read-depth and variant-allele frequency algorithms, and 3) regions of homozygosity (ROH)/uniparental disomy (UPD) analysis. Details for variant filtering and prioritization were provided in Supplement 1. Variant interpretation followed ACMG/AMP guideline^8^, with phenotype-driven prioritization of pathogenic/likely pathogenic (P/LP) variants explanatory for fetal phenotypes. Genotype-driven analysis was also used for incidental findings (IF), focusing on moderate to severe monogenic childhood-onset disorders. Reportable findings included: diagnostic P/LP variants (SNVs/Indels/CNVs/UPD), variants of uncertain significance (VUS)-high or VUS-moderate associated with phenotypes^9^, and reproductive carrier screening (RCS) results (P/LP variants only) as described by Kirk et al^10^. Secondary findings (SF) in fetus were not reported, considering the heavy burden of genetic counselling. Diagnostic yield was calculated as the proportion of cases with P/LP findings.

### CHROMOSOMAL MICROARRAY ANALYSIS

CytoScan 750K (Thermo Fisher Scientific Inc., USA) and Infinium Global/Asian Screening Array 650K (Illumina Inc., USA) were used in this study. Except Quzhou Maternal & Children Healthcare Hospital and Yancheng Maternal and Child Health Care Hospital whose 750K CMA samples were sent to Hangzhou Juno Genomics laboratories for analysis via contract service, the remaining CMA testing was performed in individual participating centers. Experimental procedures and data analysis were performed according to the instructions from the manufacturers and with their proprietary software suites. To ensure consistency on QC metrics and reporting, all CMA data were sent to Hangzhou Juno Genomics laboratories for review by a multidisciplinary team consisting of clinical geneticists, and laboratory scientists from participating institutions. Analysis identified P/LP CNVs >50 kb and clinically significant ROH (≥10 Mb or involving imprinted genes). Variant classification adhered to ACMG standards^11^.

### FOLLOW-UPS

Pregnancy outcomes were tracked via electronic health records (EHR) and physician-directed telephone follow-up for ≥3 months postpartum.

### STATISTICAL ANALYSIS

Continuous variables (maternal age, gestational age) were compared using Mann-Whitney U test; categorical variables (trimester distribution, diagnostic yield, etc.) with Chi-square test. Two-sided p<0.05 defined statistical significance. Analyses were performed with SPSS software, version 21.0 (IBM, USA).

## RESULTS

### COHORT DESCRIPTION

The study cohort comprised 1,382 singleton pregnancies: 224 in the CMA group and 1,158 in the CTW group (Figure 1, Table 1, eTable 1 in Supplement 2). After excluding 200 pregnancies terminated prior to genetic testing due to severe fetal abnormalities or multisystem anomalies (designated as the termination of pregnancy (TOP) cohort), the prospective cohort included 1,182 cases undergoing independent trio-WES and CMA testing. The median maternal age was higher in the CMA group (32 years vs 31 years; P<0.01, Table 1), in accordance with its inclusion criterion of advanced maternal age. The median maternal age was identical (31.0 years, p>0.05) for the total cohort of 1,382 cases and the prospective cohort of 1,182 cases. In contrast, the median maternal age (30.0 years) was lower in the TOP cohort compared with the total cohort (p<0.05) and with the prospective cohort (p<0.01) (eTable 2 in Supplement 2).

**Table 1.** Baseline characteristics of participants for prenatal trio exome sequencing in this study.

|  | Case number | Median | Range | *p* value |
| --- | --- | --- | --- | --- |
| Maternal age (yr) | 1382 | 31 | 17.0-47.0 | **0.006** |
| CMA group | 224 | 32 | 19.0-46.0 |  |
| CTW group | 1158 | 31 | 17.0-47.0 |  |
| Trimester | 1382 | 21.3 | 10.0-37.0 |  |
| First (1-13 wk) | 90 | 13 | 10.0-13.9 | 0.604 |
| CMA group | 3 | 13.3 | 12.7-13.3 |  |
| CTW group | 87 | 13 | 10.0-13.9 |  |
| Second (14-27 wk) | 1123 | 20.7 | 14.0-27.9 | **0.014** |
| CMA group | 207 | 19.9 | 15.0-27.9 |  |
| CTW group | 916 | 21 | 14.0-27.9 |  |
| Third (28-40 wk) | 169 | 29.7 | 28.0-37.0 | 0.124 |
| CMA group | 14 | 29.2 | 28.0-31.4 |  |
| CTW group | 155 | 29.7 | 28.0-37.0 |  |

All participants in this study were empirically divided into CMA and CTW group, respectively. CMA group included pregnant women seeking prenatal diagnosis for whom microarray analysis was traditionally recommended; CTW group included those pregnant women for whom sequential or concurrent microarray and exome sequencing were recommended.

Overall, 81.26% of participants underwent invasive procedures in the second trimester. Significant between-group differences occurred in trimester distribution (P<0.01): second-trimester referrals were more frequent in the CMA group (207/224, 92.41%) than in the CTW group (916/1,158, 79.10%), while third-trimester referrals predominated in the CTW group (155/1,158, 13.39% vs. 14/224, 6.25%; Table 1), as some structural anomalies were detected only in late pregnancy. Gestational age at diagnosis was consistently higher in the CTW group than in the CMA group for second-trimester procedures (21.0 vs 19.9 weeks, P<0.05, Table 1). The median gestational age of the total cohort was lower than that of the prospective cohort (21.3 weeks vs 21.6 weeks, p<0.05, eTable 2). Conversely, the gestational age of 20.1 weeks was substantially lower for the TOP cohort compared with the total cohort (p<0.001) and with the prospective cohort (p<0.001) (eTable 2). Follow-up data (≥3 months postpartum) were available for 1,169 pregnancies (1169/1182, 98.90%), with 13 lost to follow-up (Figure 1).

### DIAGNOSTIC YIELD OF TRIO-WES

Chromosomal abnormalities (aneuploidy/polyploidy) were detected in 2.68% (6/224) of CMA group pregnancies versus 5.87% (68/1,158; *p*=0.052) in the CTW group; clinically significant CNVs/UPD occurred in 6.25% (14/224) and 4.15% (48/1,158; *p*=0.164), respectively (Figure 2). Trio-WES identified P/LP SNVs/Indels in 4.02% (9/224) of CMA-group pregnancies— findings undetectable by CMA alone (Figure 2, eTable 3). This yield increased to 7.94% (92/1,158; *p*=0.039) in the CTW group. Of 1,382 pregnancies, 74 cases (5.35%) had chromosomal abnormalities detectable by karyotyping or CMA, while 62 cases harbored P/LP CNVs/UPD, representing a 4.49% incremental yield by CMA over karyotyping. Overall, trio-WES achieved a 7.38% (102/1,382) additional diagnostic yield (SNVs/Indels) beyond CMA, yielding a total diagnosis rate of 17.22% (238/1,382, eTable 4).

**Figure 2.**
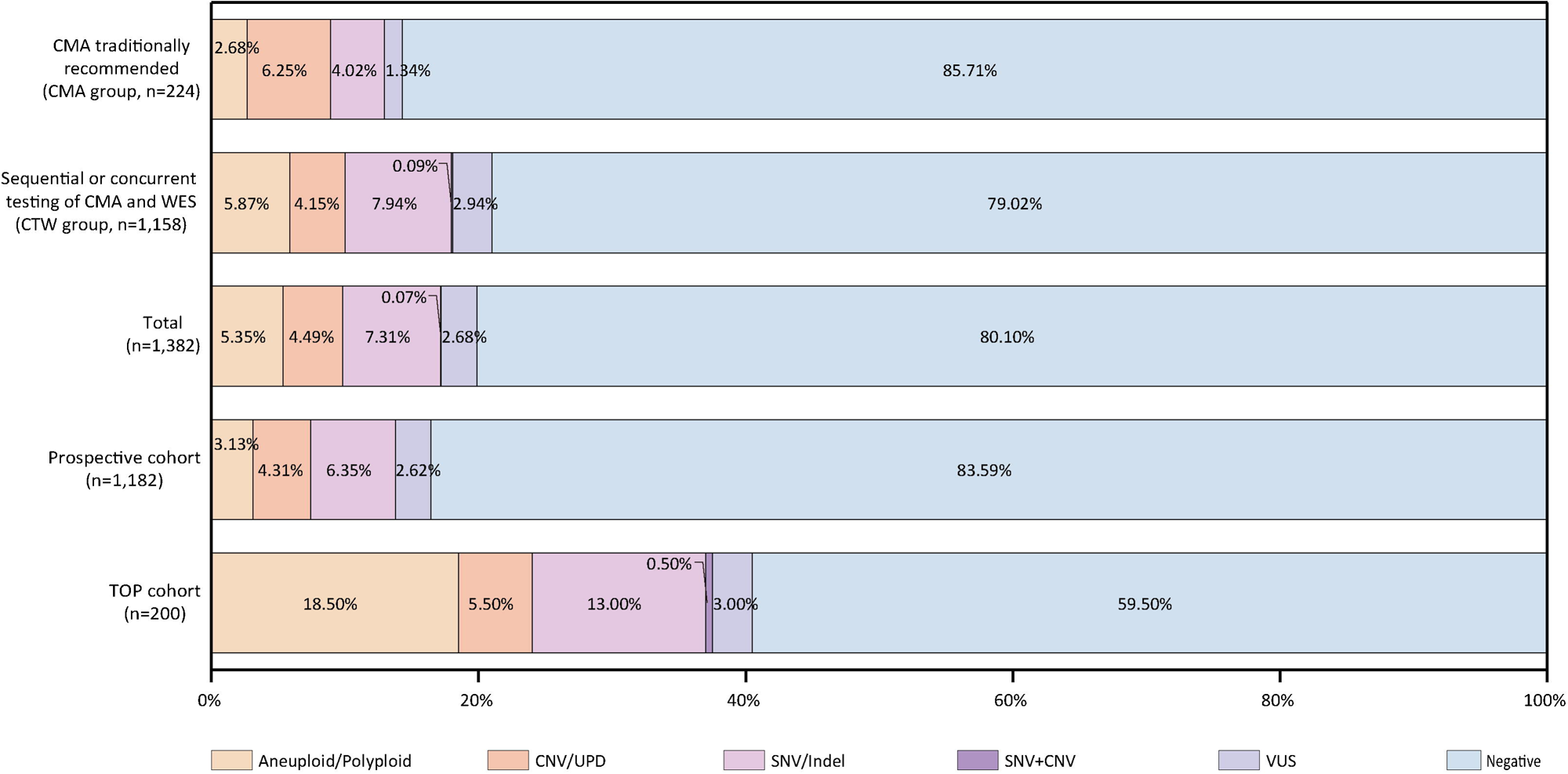
Diagnostic yield of prenatal trio exome sequencing in this study. Participants met ≥1 clinical criteria: (a) advanced maternal age, ≥35 years; (b) abnormal maternal serum biochemical screening; (c) abnormal non-invasive prenatal testing (NIPT); (d) abnormal ultrasound soft markers; (e) nuchal translucency (NT) ≥3.5mm; (f) ultrasound structural anomalies; (g) fetal growth retardation (FGR); (h) polyhydramnios/oligoamnios; (i) family history of genetic/likely genetic conditions; (j) adverse reproduction history; (k) exposure to teratogens; (l) other high risk conditions, e.g., pregnancies by in vitro fertilization (IVF) procedure. Pregnancy loss and stillbirth at enrollment were excluded from this study. Based on referral indications, subjects were stratified into two groups: 1) CMA group, in which CMA testing was traditionally recommended (standard indications: criteria a-d above), and 2) CTW group, in which CMA and WES were sequentially or concurrently recommended (expanded indications: criteria e-l). The upper row illustrates the diagnostic yield of trio-WES in the CMA group, indicating an incremental yield of 4.02% (9/224) in comparison with CMA testing; the middle raw shows the diagnostic yield of trio-WES in the CTW group with an additional yield of 7.94% (92/1,158) ; the overall diagnostic yield of 17.22% (238/1,382) of trio-WES in 1382 subjects is shown in the bottom raw with an incremental yield of 7.38% (102/1,382), compared to CMA testing alone. After excluding 200 pregnancies terminated prior to genetic testing due to severe fetal abnormalities or multisystem anomalies (designated as the TOP cohort), the prospective cohort included 1,182 cases. Significant differences in overall diagnostic yield and variant detection rates were observed between the prospective cohort and the TOP cohort. Different colors in the figure indicate different types of variations and results as illustrated at the bottom. SOC: standard-of-care; CNV: copy number variation; UPD: uniparental disomy; VUS: variant of uncertain significance; TOP: termination of pregnancy.

Significant differences in overall diagnostic yield and variant detection rates were observed between the prospective cohort and the TOP cohort. Chromosomal abnormalities were detected in 3.13% (37/1,182) of the prospective cohort versus 18.5% (37/200; p<0.001) of the TOP cohort. CNVs/UPDs occurred in 4.31% (51/1,182) versus 5.5% (11/200; p>0.05), and SNVs/Indels in 6.35% (75/1,182) versus 13.0% (26/200; p<0.001). The overall trio-WES diagnostic yield was 13.79% in the prospective cohort and 37.5% in the TOP cohort (p<0.001) (Figure 2, eTable 5, eFigure 2).

### CYTOGENETIC EVENTS BY CMA VERSUS TRIO-WES

In the prospective cohort (n=1,182), 41 cases (3.49%) were excluded from concordance analysis due to inadequate DNA quality (n=26) or quantity (n=15). Among 1,141 analyzable cases, trio-WES demonstrated complete concordance with CMA for detecting all reportable cytogenetic events, including chromosomal abnormalities and CNVs/UPD (Figure 3, eTable 6). Trio-WES further resolved inheritance patterns for 29 CMA-identified VUS, enabling refined clinical interpretation. Crucially, trio-WES detected P/LP SNVs/indels in 5.96% (68/1,141) of pregnancies with negative or inconclusive CMA results (Figure 3, eTable 7). Analysis of 17 CMA-reported ROH revealed six cases of UPD (6/1141, 0.53%), providing key information for risk assessment of imprinting disorders. Additionally, trio-WES identified clinically significant exon-level deletions in 0.26% (3/1,141) of cases not detected by CMA (eTable 8). In sum, trio-WES achieved an additional 8.85% of informative results compared to CMA testing, including 2.1% case with VUS SNVs/Indels.

**Figure 3.**
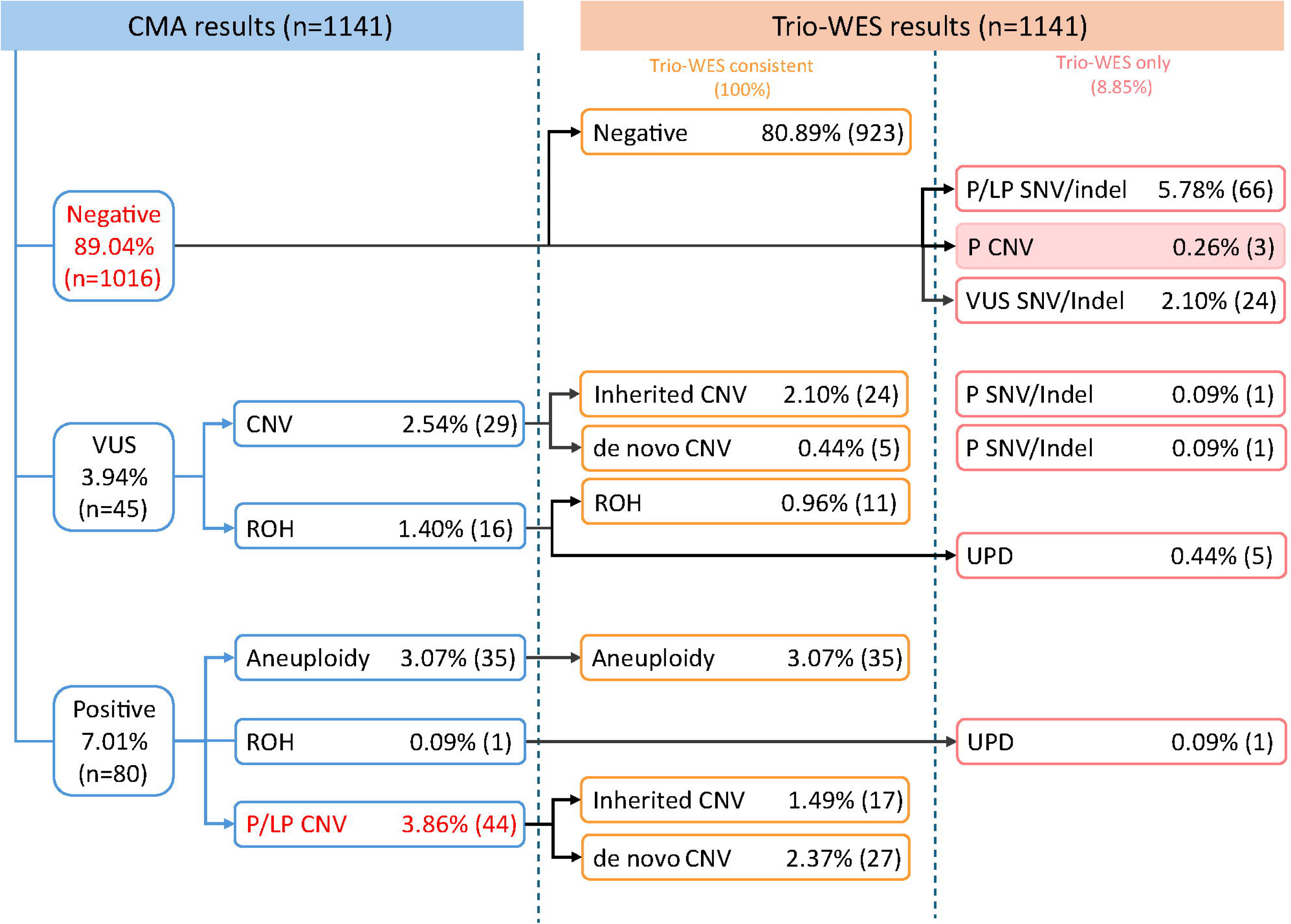
Comparison on prenatal detection of genetic variants by trio-WES and CMA in this study. Of 1182 pregnancies in the prospective cohort, 1141 cases underwent independent trio-WES and CMA testing in parallel and were used to comparison on detection of genetic variants between these two different testing approaches, with 26 cases of DNA QC failure and 15 cases of inadequate DNA quantity. The CMA results of 1141 cases were shown in the left column, the consistent results of both CMA and trio-WES in the middle column, and the trio-WES only results in the right. An additional informative result of 8.85% (101/1141) was obtained by trio-WES that would be missed by CMA, with a consistency of 100% on detection of clinically reportable CNVs. VUS: variant of uncertain significance; CNV: copy number variation; ROH: run of homozygosity; P/LP: pathogenic/likely pathogenic; UPD: uniparental disomy

### CLINICAL UTILITY OF PRENATAL TRIO-WES

Among 1,169 cases with complete follow-up data, 162 had positive results (eTable 9). Among 120 pregnancies with *de novo*, compound heterozygous, homozygous or hemizygous variants, the rate of TOP were 90.28% (65/72) for cytogenetic events and 75.0% (36/48) for SNVs/Indels, respectively. In contrast, TOP rates were lower for inherited variants: 25.0% (4/16) for cytogenetic events and 42.31% (11/26) for SNVs/Indels. These differences were statistically significant for cytogenetic events (90.28% vs. 25.0%; P<0.001), SNVs/Indels (75.0% vs. 42.31%; P<0.01), and combined variants (84.17% [101/120] vs. 35.71% [15/42]; P<0.001) (Figure 4, eTable 10). Consequently, parents of fetuses with parental inherited variants were significantly more likely to continue the pregnancy to live birth than parents of fetuses with *de novo*, biallelic or hemizygous variants (27/42, 64.29% vs. 19/120, 15.83%; P<0.001) (Figure 4).

**Figure 4.**
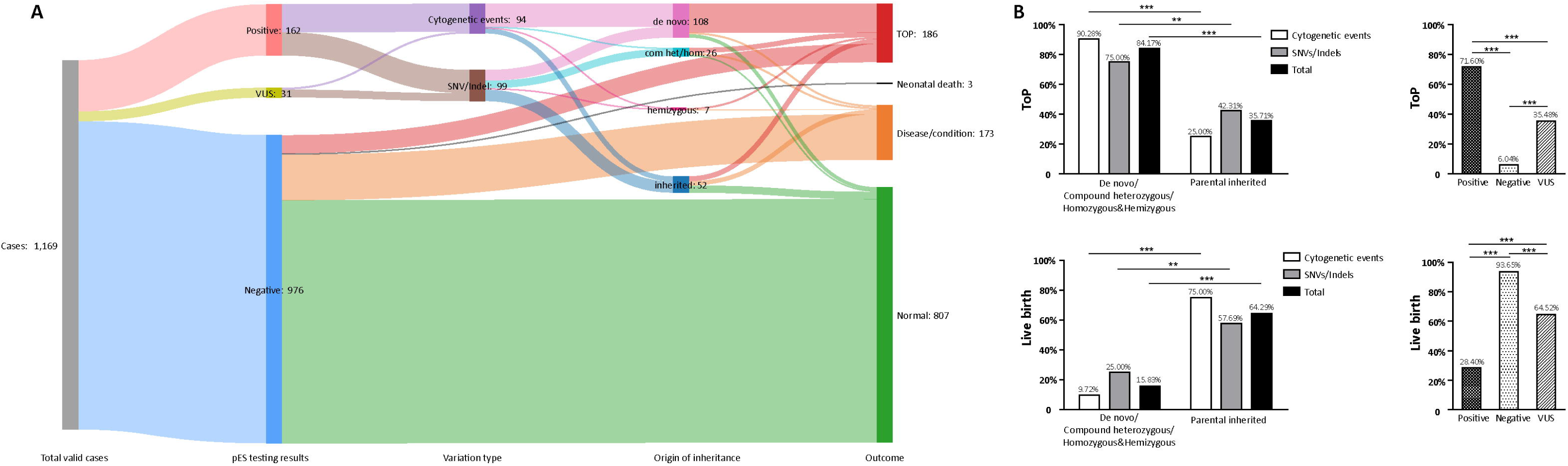
Outcomes and the impact of trio-WES testing on clinical decision-making for pregnancies in this prospective cohort. Sankey diagram of pregnancy outcomes according to trio-WES testing results in 1169 cases in whom a follow-up of at least 3 months postpartum was obtained from this prospective multi-center cohort of 1182 pregnancies. The thickness is proportional to the number of cases in every category as depicted, and the color represents individual status as described (A). For pregnancies with positive trio-WES results (either cytogenetic events or SNVs/Indels or both combined), the rate of TOP (upper left) and live birth (down left) between fetuses harboring *de novo* or biallelic or hemizygous variants and fetuses with parental inherited variants differed significantly. Similarly, the rate of TOP (upper right) and live birth (down right) among pregnancies with positive, negative, and inconclusive (VUS) trio-WES results also significantly differed in this study, demonstrating the important impact of genetic testing on clinical decision-making for pregnancies seeking invasive prenatal diagnosis (B). **denotes p<0.01; ***denotes p<0.001; VUS: variant of uncertain significance; com het: compound heterozygous; hom: homozygous; TOP: termination of pregnancy.

For another, 976 cases (976/1,169, 83.49%) yielded negative results. Of these, 769 pregnancies (78.79%) resulted in live births with normal developmental milestones during postnatal follow-up (≥3 months). TOP occurred in 59 cases (6.05%), and neonatal death was documented in 3 cases (0.31%) (eTable 11). The remaining 145 pregnancies (14.86%) resulted in live births with disease manifestations, including persistent fetal phenotypes or phenotypes newly emerged in the neonatal period. Further analysis revealed that clinical decision-making was jointly influenced by severity of the fetal clinical phenotypes, perceived fetal prognosis, and the molecular diagnostic findings. For instance, pregnancies involving isolated congenital heart defects (e.g., ventricular septal defect) with negative trio-WES results, representing 72 of 145 disease-associated live births (49.66%), predominantly continued pregnancy. Conversely, pregnancies with complex congenital heart disease (16.95%) or multi-system anomalies (38.98%) more frequently resulted in TOP (eFigure 3).

Reanalysis of trio-WES data was performed for 145 fetuses with initially negative results but postnatal disease phenotypes. Clinically relevant variants explaining the observed phenotypes were identified in 10 cases (6.90%, eTable 12): thalassemia minor (n=6), ichthyosis vulgaris (n=2), glucose-6-phosphate dehydrogenase deficiency (n=1), and overgrowth syndrome (n=1). Among these, nine variants (excluding overgrowth syndrome) corresponded to conditions without observable prenatal phenotypes. Consistent with the predefined reporting policy for prenatal WES, these nine variants were not originally reported due to mild phenotypic severity or limited clinical actionability.

Additionally, 31 fetuses (31/1169, 2.65%) carried VUS (eTable 13). Of these 31 pregnancies, TOP occurred in 11 cases (35.48%) following genetic counseling; all TOP cases involved *de novo*, compound heterozygous, homozygous, or hemizygous variants. The TOP rate for VUS carriers was significantly lower than that for cases with P/LP variants (35.48% vs. 71.60%; P<0.001; Figure 4) but higher than for negative cases (35.48% vs. 6.04%; P<0.001). Live births with normal postnatal phenotypes were documented in 12 cases (38.71%). The remaining 8 pregnancies (25.81%) resulted in live births with persistent or progressive prenatal abnormalities, prompting data reanalysis. This reanalysis identified 2 variants reclassified as P/LP, explaining the clinical presentations (hypothyroidism in one case and Mowat-Wilson syndrome in another). Six variants retained VUS classification, 4 of which were considered potentially explanatory for the observed phenotypes (eTable 14).

### RCS

This study applied RCS targeting early-onset or lethal disorders as defined by Kirk et al^10^. In the prospective cohort of 1,182 couples, 37 (3.13%) met high-risk criteria (eTable 15), a proportion lower than that reported by Kirk et al. Seventeen high-risk couples (1.44%) were newly identified through RCS, while 20 couples (1.69%) had prior knowledge of carrier status, including three couples who underwent IVF. Prenatal testing identified compound heterozygous, homozygous, or hemizygous variants inherited from high-risk parents in 11 of 37 fetuses. TOP occurred in 8 of these 11 cases (72.73%).

## DISCUSSION

The diagnostic yield of prenatal trio-WES in this cohort was 17.22%. After excluding cases with cytogenetic abnormalities primarily detectable by conventional karyotyping and CMA, the yield was 7.38%. This figure is modestly lower than the 8.5% reported in the PAGE study^2^ and 10% in study by Petrovski et al^3^, and significantly below the 12.44% observed by Liao et al^12^. These discrepancies primarily reflect divergent inclusion criteria across studies. Whereas the PAGE and Petrovski’s studies exclusively enrolled fetuses with structural anomalies, and Liao et al. restricted inclusion to cases with NT ≥3.5 mm, structural anomalies, or hydrops fetalis, our cohort encompassed a much broader phenotypic spectrum involving diverse risks. The lower diagnostic rate in the prospective cohort (13.79% vs. 17.22% overall) may be partially attributed to the exclusion of 200 cases. Notably, the TOP cohort demonstrated a substantially higher diagnostic yield of 37.5%, with 13.0% of cases harboring P/LP SNVs/Indels beyond chromosomal abnormalities and CNVs. These findings support the clinical utility of trio-WES in pregnancies with severe adverse outcomes, provide benchmark data for diverse prenatal populations, and highlight cases of the TOP cohort as a high-priority subgroup for comprehensive genetic assessment to inform reproductive counseling.

To address the diagnostic ambiguity of “soft” indications, we prospectively stratified pregnancies with advanced maternal age (≥35 years), abnormal serum biochemical screening, NIPT anomalies, or isolated soft markers into a dedicated CMA subgroup. Within this group, trio-WES provided an incremental diagnostic yield of 4.02% beyond CMA. This exceeds the yields reported by Levy et al. (2.7% in low-risk pregnancies)^5^ and Vaknin et al. (2.86% in structurally normal fetuses)^4^. Critically, our CMA group cohort, which was enriched for high-risk biochemical/NIPT profiles, differs fundamentally from the low-risk populations in these comparator studies, explaining the observed variance. These findings provide solid evidence for trio-WES as the first-tier genetic testing for all pregnancies seeking prenatal diagnosis.

Whereas CMA remains the diagnostic cornerstone for invasive prenatal testing in resource-limited settings, our prospective cohort study of 1,141 pregnancies demonstrates that trio-WES achieves complete concordance with CMA in detecting clinically significant CNVs, while simultaneously resolving critical limitations of conventional approaches. Although Danecek et al. previously established WES-based CNV detection’s superiority over low-resolution CMA platforms (e.g., CytoScan 750K) and parity with exon-resolution arrays^7^, our study provides the first rigorous validation in clinical prenatal diagnostics. Beyond equivalence in CNV identification, trio-WES delivers transformative advantages, including precisely determining genetic origins of variants to reduce ambiguous VUS reports, detecting exonic CNVs inaccessible to CMA, and enabling UPD identification essential for diagnosing imprinting disorders. Consequently, our data position trio-WES not merely as an adjunct test but as the foundational platform for prenatal genomic medicine, redefining standards of care for global implementation regardless of resource constraints.

Parental decisions integrate multifactorial considerations including disease severity, prognosis, and therapeutic availability alongside genetic results. Our findings demonstrate that the classification and inheritance patterns of P/LP variants significantly influence clinical decision-making, evidenced by much higher termination rates for *de novo* or biallelic P/LP variants (84.17%) versus inherited P/LP variants (35.71%). This pattern strongly supports the emerging understanding that certain genetic findings may be perceived as more manageable to intervention, thereby altering the threshold for pregnancy continuation – a concept reinforced by the comprehensive phenotypic and genotypic insights afforded by trio-WES. Critically, even in the absence of a definitive molecular diagnosis (WES-negative), the application of prenatal trio-WES remains an indispensable component of the diagnostic workup, providing crucial negative predictive information and facilitating the exclusion of a vast spectrum of monogenic disorders, thereby informing clinical management with a comparable scenario where a positive diagnosis is established. Additionally, data reanalysis provided substantial added value: VUS reclassification resolved diagnostic uncertainties in two families, while retrospective analysis yielded diagnoses in 6.9% (10/145) of initially negative cases with postnatal phenotypes. Nevertheless, the effective implementation of systematic data reanalysis, particularly in the context of prenatal WES, faces substantial technical and interpretive challenges that warrant dedicated future research^13, 14^.

Current SF frameworks^15^ focus primarily on postnatal care, demonstrating limited relevance to prenatal diagnosis through minimal gene list overlap (8/81 genes) with fetal treatable genes proposed by Cohen et al.^16^, a clinically actionable set of 296 genes targeting conditions amenable to in utero or immediate postnatal intervention. Our study did not address fetal-specific SF curation - a critical gap requiring multidisciplinary consensus to establish evidence-based prenatal SF guidelines. Additional limitations include the modest CMA cohort size, necessitating larger studies to define trio-WES utility in this population, and constrained follow-up duration that precluded assessment of developmental phenotypes. Longitudinal phenotyping initiatives are essential to refine variant interpretation across prenatal testing modalities, including emerging approaches like whole-genome sequencing (WGS) and transcriptomic profiling (RNA-Seq).

Although WGS offers enhanced detection of noncoding variants and structural rearrangements^17^, its diagnostic utility remains constrained by the frequent need for corroborating clinical or familial segregation data. Concurrently, WES capabilities have expanded through optimized probe design and bioinformatic pipelines, now encompassing exon-level CNVs, mitochondrial variants, select repeat expansions, and known noncoding variants—narrowing its performance gap with WGS. Given its favorable cost-effectiveness and reduced analytical complexity, WES remains the pragmatic first-tier prenatal diagnostic modality, particularly in resource-limited settings. WGS implementation may advance with further technological refinements and fetal-specific interpretation frameworks.

To our knowledge, this represents the largest prospective multicenter study conducted to date that rigorously assesses the diagnostic yield of trio-WES across a broad spectrum of prenatal indications extending beyond isolated structural anomalies, systematically evaluates the performance of WES-based CNV detection in the prenatal setting, and comprehensively examines the clinical utility of trio-WES through longitudinal follow-up. Our findings provide a robust evidence base supporting the adoption of trio-WES as a stand-alone, first-tier diagnostic approach for pregnancies undergoing invasive prenatal diagnosis.

## Data Availability

All data produced in the present study are available upon reasonable request to the authors

## Funding

This work was supported by grants from Ningbo science and technology project (No.2023Z178), Innovation Project of Distinguished Medical Team in Ningbo (No.2022020405), both to Changshui Chen; the Social Development Public Welfare Foundation of Ningbo (No.2022S035), Key Technology Breakthrough Program of Ningbo Sci-Tech Innovation Yongjiang 2035 (No.2024Z221), both to Haibo Li; and Ningbo Natural Science Foundation Project (2024J358) to Lulu Yan.

## Authors’ contributions

HBL, ZJZ, and JG conceptualized the study. ML, LYT, JC, YLD, YYZ, HMZ, SSS, GSS, PLC, SSW, XPS, and XYX performed the cohort recruitment, follow-ups, pre-, and post-test genetic counseling. LLY, JYX, YXZ, ML, GSS, PLC, YYZ, YLD, XPS, SF and XYX collected samples and relevant clinical data, and processed the clinical data. HBL, LLY, and CSC acquired the funding. LLY, JYX, YXZ, GSS, PLC, XPS, YLD, and XYX performed the wet experiments. YCW and DJL performed the bioinformatic analysis. HBL, LLY, JYX, YXZ, ML, YLD, XPS, PLC, SSS, HMZ, DDC, and JG performed medical interpretations. HBL, ML, YTX, JG, and ZJZ did the formal analysis. YTX, GJ, and ZJZ performed the statistics. HBL, ZJZ, and JG wrote the original draft. HBL, JG, ZJZ, and CSC reviewed and edited the manuscript. HBL, GJ, ZJZ, and CSC supervised the whole process of research. HBL, SF, and CSC performed the project coordination. All authors read and approved the final manuscript.

## Acknowledgements

We are deeply grateful to the study participants and their families. We also thank the personnel and administrative staff at the seven participating institutions for their essential work in subject enrollment, sample collection, and follow-up. We are particularly indebted to the staff of the Division of Bioinformatics, Core Facility of High-Throughput Sequencing, and Department of Medical Affairs at Hangzhou Juno Genomics Inc. for their vital contributions. Finally, we thank Mr. Xiao Li (The X Laboratory and Center for Mendelian Genomics, Advanced Institute of Information Technology, Peking University) for his expert assistance in bioinformatic analysis.

Dr. Juan Geng is a co-founder of and consulted for Hangzhou Juno Genomics Inc. Dr. Zhaojing Zheng is a co-founder and chief executive officer of Hangzhou Juno Genomics Inc. Yanting Xu, Dandan Cai, Yuchao Wang, Danjun Liang, and Shu Fang are employees of Hangzhou Juno Genomics Inc. All other authors declare no potential conflicts of interest.

## Notes

### Funding Statement

This study was funded by Ningbo science and technology project (No.2023Z178)
Innovation Project of Distinguished Medical Team in Ningbo (No.2022020405)
Social Development Public Welfare Foundation of Ningbo (No.2022S035)
Key Technology Breakthrough Program of Ningbo Sci-Tech Innovation Yongjiang 2035 (No.2024Z221)
Ningbo Natural Science Foundation Project (2024J358) to Lulu Yan.

### Author Declarations

This study was approved by the Ethics Committee of The Affiliated Women and Children's Hospital of Ningbo University (EC2020-014), Quzhou Maternity & Child Healthcare Hospital (EC-KY2023-03), Lishui Maternity & Child Healthcare Hospital (2020034), Jinhua Maternity & Child Healthcare Hospital (EC2021-KY030), Huzhou Maternity & Child Healthcare Hospital (2020-R-029), Yancheng Maternity & Child Healthcare Hospital (2022-NT-005), The First Affiliated Hospital of Chongqing Medical University (K2023-580).

## Reference

1. Wapner RJ, Martin CL, Levy B, et al. Chromosomal Microarray versus Karyotyping for Prenatal Diagnosis. N Engl J Med. 2012;367(23):2175–2184. doi:10.1056/NEJMoa1203382

2. Lord J, McMullan DJ, Eberhardt RY, et al. Prenatal exome sequencing analysis in fetal structural anomalies detected by ultrasonography (PAGE): a cohort study. The Lancet. 2019;393(10173):747–757. doi:10.1016/s0140-6736(18)31940-8

3. Petrovski S, Aggarwal V, Giordano JL, et al. Whole-exome sequencing in the evaluation of fetal structural anomalies: a prospective cohort study. The Lancet. 2019;393(10173):758–767. doi:10.1016/s0140-6736(18)32042-7

4. Vaknin N, Azoulay N, Tsur E, et al. High rate of abnormal findings in Prenatal Exome Trio in low risk pregnancies and apparently normal fetuses. Prenat Diagn. 2021;42(6):725–735. doi:10.1002/pd.6077

5. Levy M, Lifshitz S, GoldenbergDFumanov M, et al. Exome sequencing in every pregnancy? Results of trio exome sequencing in structurally normal fetuses. Prenat Diagn. 2024;45(3):276–286. doi:10.1002/pd.6585

6. Sparks TN. Prenatal exome sequencing for the structurally normal fetus: ready or not? Am J Obstet Gynecol MFM. 2025;7(2)doi:10.1016/j.ajogmf.2024.101513

7. Danecek P, Gardner EJ, Fitzgerald TW, et al. Detection and characterization of copy-number variants from exome sequencing in the DDD study. Genet Med Open. 2024;2doi:10.1016/j.gimo.2024.101818

8. Richards S, Aziz N, Bale S, et al. Standards and guidelines for the interpretation of sequence variants: a joint consensus recommendation of the American College of Medical Genetics and Genomics and the Association for Molecular Pathology. Genet Med. 2015;17(5):405–424. doi:10.1038/gim.2015.30

9. Bennett G, Karbassi I, Chen W, et al. Distinct rates of VUS reclassification are observed when subclassifying VUS by evidence level. Genet Med. 2025;27(6)doi:10.1016/j.gim.2025.101400

10. Kirk EP, Delatycki MB, Archibald AD, et al. Nationwide, Couple-Based Genetic Carrier Screening. N Engl J Med. 2024;391(20):1877–1889. doi:10.1056/NEJMoa2314768

11. Riggs ER, Andersen EF, Cherry AM, et al. Technical standards for the interpretation and reporting of constitutional copy-number variants: a joint consensus recommendation of the American College of Medical Genetics and Genomics (ACMG) and the Clinical Genome Resource (ClinGen). Genet Med. 2020;22(2):245–257. doi:10.1038/s41436-019-0686-8

12. Fu F, Li R, Yu Q, et al. Application of exome sequencing for prenatal diagnosis of fetal structural anomalies: clinical experience and lessons learned from a cohort of 1618 fetuses. Genome Med. 2022;14(1)doi:10.1186/s13073-022-01130-x

13. Thauvin-Robinet C, Garde A, Favier M, et al. Reanalysis of unsolved prenatal exome sequencing for structural defects: diagnostic yield and contribution of postnatal/postmortem features. Eur J Hum Genet. 2025;33(5):675–682. doi:10.1038/s41431-025-01823-y

14. Mone F, Abu Subieh H, Doyle S, et al. Evolving fetal phenotypes and clinical impact of progressive prenatal exome sequencing pathways: cohort study. Ultrasound Obstet Gynecol. 2022;59(6):723–730. doi:10.1002/uog.24842

15. Miller DT, Lee K, Abul-Husn NS, et al. ACMG SF v3.2 list for reporting of secondary findings in clinical exome and genome sequencing: A policy statement of the American College of Medical Genetics and Genomics (ACMG). Genet Med. 2023;25(8)doi:10.1016/j.gim.2023.100866

16. Cohen JL, Duyzend M, Adelson SM, et al. Advancing precision care in pregnancy through a treatable fetal findings list. Am J Hum Genet. 2025;112(6):1251–1269. doi:10.1016/j.ajhg.2025.03.011

17. Wojcik MH, Lemire G, Berger E, et al. Genome Sequencing for Diagnosing Rare Diseases. N Engl J Med. 2024;390(21):1985–1997. doi:10.1056/NEJMoa2314761

