## Supplement 1 for "Trio exome sequencing as the stand-alone first-tier testing for prenatal diagnosis"

**Methods**

**Subject recruitment**

In this study we enrolled singleton pregnant women who were offered invasive prenatal diagnosis due to high risk for fetus anomaly during their prenatal visits to the prenatal diagnosis center from the seven hospitals included in this multi-center prospective study. The seven centers came from The Affiliated Women and Children's Hospital of Ningbo University, Quzhou Maternity & Child Healthcare Hospital, Lishui Maternity & Child Healthcare Hospital, Jinhua Maternity & Child Healthcare Hospital, Huzhou Maternity & Child Healthcare Hospital, all in Zhejiang Province; Yancheng Maternity & Child Health care Hospital, Jiangsu Province; The First Affiliated Hospital of Chongqing Medical University, Chongqing. Recruitment was completed via the ordering physicians from the seven prenatal diagnosis centers from Oct 1, 2022 to April 30, 2024. Due to variability in administration efficiency and regulation policy in different hospitals, the sampling date and sample size differed accordingly. In addition to prenatal WES, the seven centers routinely also provide other standard-of-care (SOC) genetic tests including CMA and karyotyping. The ordering physicians may offer some patients SOC tests concurrently or sequentially with WES whenever they thought appropriate.

Subjects included in this study met at least one of the following categories: (a) advanced maternal age, ≥35 years; (b) abnormal maternal serum biochemical screening; (c) abnormal non-invasive prenatal testing (NIPT); (d) abnormal ultrasound soft markers; (e) nuchal translucency (NT) ≥3.5mm; (f) ultrasound structural anomalies; (g) fetal growth retardation (FGR); (h) polyhydramnios/oligoamnios; (i) family history of genetic/likely genetic conditions; (j) adverse reproduction history; (k) exposure to teratogens; (l) other high risk conditions, e.g., pregnancies by in vitro fertilization (IVF) procedure. Low-risk structural normal pregnancies and non-singleton pregnancies were not enrolled in this study. According to the referral reasons, subjects were stratified into two groups: 1) CMA group, in which CMA testing was traditionally recommended (standard indications: criteria a-d above), and 2) CTW group, in which CMA and WES were sequentially or concurrently recommended (expanded indications: criteria e-l).

Written consent was obtained from all subjects after pre-test counselling by the ordering physician. This study was approved by the Ethics Committee of The Affiliated Women and Children's Hospital of Ningbo University (EC2020-014), Quzhou Maternity & Child Healthcare Hospital (EC-KY2023-03), Lishui Maternity & Child Healthcare Hospital (2020034), Jinhua Maternity & Child Healthcare Hospital (EC2021-KY030), Huzhou Maternity & Child Healthcare Hospital (2020-R-029), Yancheng Maternity & Child Healthcare Hospital (2022-NT-005), The First Affiliated Hospital of Chongqing Medical University (K2023-580).

**Genomic DNA extraction**

The genomic DNA (gDNA) from amniotic fluid (AF), cultured amniocytes, chorionic villus sampling (CVS), fetal tissue, umbilical cord blood, peripheral blood was extracted on Concert Bio HF48 Nuclear Acid Extractor system with Genomic DNA Tissue Extraction kit, Genomic DNA Cultured Cell Extraction kit, and Genomic DNA Whole Blood Extraction kit, respectively (all from Concert Bioscience (Xiamen) Co.,Ltd., China). Sample quality control (QC) was performed with Qubit Flex Fluorometer and NanoDrop Microvolume Spectrophotometers (Thermo Fisher Scientific Inc., USA). Maternal cell contamination (MCC) was measured with AneuFiler^TM^ kit (Shanghai Cubicise Medical Co., Ltd., China), a QF-PCR assay designed for detection of aneuploidy of chromosome 13, 18, 21, and sex chromosomes. Data was generated by Applied Biosystems 3500 Dx Genetic Analyzer and analyzed with GeneMapper software v6.0 (Thermo Fisher Scientific Inc., USA) for all fetal gDNA. Samples with MCC level ≧20% were excluded from further testing in this study.

**Whole exome sequencing**

A central-lab approach was applied in this study. WES experiments were performed in laboratories of Hangzhou Juno Genomics Inc. DNA libraries were constructed using the Library Preparation EF 2.0 Kit with Enzymatic Fragmentation and the Twist Universal Adapter System (Twist Bioscience, USA). gDNA was enzymatically fragmented, followed by end repair, dA-tailing, adapter ligation, and PCR amplification. Custom exome capture was performed using the OmniSeek^®^ Whole Exome Sequencing panel (Hangzhou Juno Genomics Inc., China) and Twist Fast Hybridization and Wash Kit according to the manufacturer’s instructions. The Custom OmniSeek^®^ Exome panel consists of the Twist Comprehensive Exome Panel (Cat. No.:102033), the Twist Mitochondrial Panel Kit (Cat. No.:102040), and Custom Spike-in Panel TE-97665586（Cat. No.:101282), which included probes against known clinically relevant non-coding P/LP variants, probes against known clinically significant microdeletions and microduplications, and Genome-wide CNV Backbone probes. The Custom OmniSeek^®^ Exome panel was comprehensively optimized and validated with NA12878 samples to ensure consistent and high-quality coverage of region of interest (ROI). Subsequently, the captured libraries were sequenced on the DNBSEQ-T7 platform (MGI Tech Co., Ltd., China) with 150 bp paired-end reads and a mean depth of at least 100X.

**Bioinformatic analysis**

All bioinformatic analytic works were completed via the cloud platform JunoCloud^®^ developed by Hangzhou Juno Genomics Inc. Briefly, raw FASTQ reads were preprocessed using fastp ^1^ (v0.23.1) to filter out low-quality and adapter-contaminated reads, resulting in clean reads which were subsequently aligned to the human reference genome (GRCh38/Hg38) using the BWA ^2^ (v0.7.17-r1188) mem algorithm. Sambamba ^3^ (v0.8.2) was used to convert alignments to BAM format and remove PCR duplicates.

Variant calling was performed with the Genome Analysis Toolkit (GATK) ^4^ (v4.2.4.0), utilizing the HaplotypeCaller workflow. Variant annotation was carried out using Ensemble’s Variant Effect Predictor (VEP) ^5^ (v105) and ANNOVAR ^6^ (v2020Jun.08). Annotation was completed with ANNOVAR and multiple reference datasets, including Online Mendelian Inheritance in Man (OMIM), Orphanet, ClinVar, ClinGen, 1000G Phase 3 v5a, the Genome Aggregation Database (gnomAD v4.1), the Exome Aggregation Consortium (ExAC 3.0), the Exome Sequencing Project (ESP6500siv2_all), the professional versions of the Human Gene Mutation Database (HGMD professional v2018.2 & v2021.2), and an in-house Han Chinese patients population frequency database JunoDB^®^. Furthermore, several functional prediction tools were employed, including LJB* (dbNSFP) ^7,^ avSNP150^8^, REVEL ^9^ and ClinPred ^10^, for protein function predictions, while SCAP ^11^ and SpliceAI ^12^ was used to assess potential splicing impacts.

Two approaches were used to determine the sample's gender and the family member's relationship, i.e., the average depth of specific genes on chromosome Y and the heterozygous variants percentage on chromosome X. A custom algorithm was developed to analyze multiple single nucleotide polymorphisms (SNPs) for inheritance patterns across family members, aiming to confirm family pedigree relationships.

Moreover, CNV calling from WES data was carried out using three independent tools: XHMM ^13^, CNVkit ^14^ (v0.9.10), and DECoN ^15^ (v1.0.2). Runs of homozygosity (ROH) were detected using H3M2Tool ^16^ and AutoMap ^17^ (v1.0). Uniparental disomy (UPD) analysis was performed using UPDio ^18^ (v1.0). Repeat expansions were assessed using ExpansionHunter ^19^ (v5.0.0). Mobile element insertions were identified using SCRAMble ^20^ (v1.0.2).

The QC metrics for the data included the following: (1) the average data output per case was ≥12 Gb; (2) the average depth for core exome regions ≥100X, with ≥98% of these regions achieving at least 20X coverage; (3) the average depth for mitochondrial genome was ≥2000X.

**Variant filtering and prioritization**

SNVs were excluded from downstream analysis based on the following quality criteria: (1) Quality by Depth (QD) < 2.0; (2) Fisher Strand (FS) > 60.0; (3) Strand Odds Ratio (SOR) > 3.0; (4) Mapping Quality (MQ) < 30; (5) Mapping Quality Rank Sum (MQRankSum) < -12.5; (6) Read Pos Rank Sum (ReadPosRankSum) < -8.0. For insertion/deletion variants (INDELs), more stringent filtering criteria were applied: (1) QD < 2.0; (2) FS > 200.0; (3) SOR > 10.0; (4) MQRankSum < -12.5; (5) ReadPosRankSum < -8.0.

All QC-pass annotated variants were subjected to downstream analysis with the in-house scripts. Variants with a minor allele frequency (MAF) > 5% were excluded, except for those listed in the HGMD, ClinVar, and ClinGen BA1 exception list ^21^ (BA1). To facilitate the interpretation of variants, phenotype information for each fetus was extracted from the test requisition forms (TRFs) and converted into the standard Human Phenotype Ontology (HPO) terms.

A genotype-driven approach was employed to prioritize rare variant lists for each fetus. Variants with an allele frequency of less than 0.1% were specifically selected for further analysis. The prioritization strategy included the following categories: (1) dominant *de novo* variants; (2) recessive homozygous variants, excluding those identified in homozygous status in internal healthy control datasets; (3) recessive compound heterozygous variants; (4) *de novo* X chromosome variants or rare hemizygous variants inherited from mother; (5) known pathogenic alleles, excluding those classified as benign or likely benign in ClinVar; and (6) predicted truncating variants (nonsense, frameshift, canonical splice site alterations). The resulting gene/variant list was subsequently assessed for clinical relevance, with potentially significant variants categorized according to the American College of Medical Genetics and Genomics (ACMG) guidelines ^22^ and ClinGen Variant Curation Expert Panels (VCEP) gene-specific criteria ^23-30^, as applicable.

Secondary findings (SF) in fetus were not reported in this study due to heavy burden of genetic counselling and potential psychological impact on parents after achieving a consensus on this issue by ordering physicians from the seven participating centers. Incidental findings (IF) were restricted to genes associated with early-onset diseases or diseases for which clinical intervention measures are available.

**Pathogenicity assessment**

The classification and assessment of pathogenicity of variants was performed according to ACMG/AMP guidelines ^22^. Only P/LP variants were reported and used for comparison throughout this study.

**Tools**

1. Fastp: https://github.com/OpenGene/fastp
2. BWA: https://bio-bwa.sourceforge.net/
3. Sambamba: https://github.com/biod/sambamba
4. Genome Analysis ToolKit: https://gatk.broadinstitute.org/hc/en-us
5. Ensembl’s Variant Effect Predictor: https://www.ensembl.org/info/docs/tools/vep/index.html
6. Annovar: https://annovar.openbioinformatics.org/en/latest/
7. dbNSFP: https://www.dbnsfp.org/
8. avSNP150: https://www.ncbi.nlm.nih.gov/snp/
9. REVEL: https://sites.google.com/site/revelgenomics/
10. ClinPred: https://sites.google.com/site/clinpred/
11. SCAP: http://bejerano.stanford.edu/scap/
12. SpliceAI: https://github.com/Illumina/SpliceAI
13. XHMM: https://github.com/statgen/XHMM
14. CNVkit: https://github.com/etal/cnvkit
15. DECoN: https://github.com/RahmanTeam/DECoN
16. H3M2Tool: https://sourceforge.net/projects/h3m2/
17. AutoMap: https://github.com/mquinodo/AutoMap
18. UPDio: https://github.com/findingdan/UPDio
19. ExpansionHunter: https://github.com/Illumina/ExpansionHunter
20. SCRAMble: <https://github.com/GeneDx/scramble>

**Resources**

OMIM: https://omim.org/

Orphanet: https://www.orpha.net/

ClinVar: https://www.ncbi.nlm.nih.gov/clinvar/

ClinGen: https://clinicalgenome.org/

1000G Phase 3: https://www.internationalgenome.org/data-portal/data-collection/phase-3

gnomAD: https://gnomad.broadinstitute.org/news/2024-04-gnomad-v4-1/

HGMD: https://www.hgmd.cf.ac.uk/ac/index.php

HPO: https://hpo.jax.org/

**References**

1.	Chen S, Zhou Y, Chen Y, Gu J. fastp: an ultra-fast all-in-one FASTQ preprocessor. *Bioinformatics*. Sep 1 2018;34(17):i884-i890. doi:10.1093/bioinformatics/bty560

2.	Li H, Durbin R. Fast and accurate short read alignment with Burrows-Wheeler transform. *Bioinformatics*. Jul 15 2009;25(14):1754-60. doi:10.1093/bioinformatics/btp324

3.	Tarasov A, Vilella AJ, Cuppen E, Nijman IJ, Prins P. Sambamba: fast processing of NGS alignment formats. *Bioinformatics*. Jun 15 2015;31(12):2032-4. doi:10.1093/bioinformatics/btv098

4.	McKenna A, Hanna M, Banks E, et al. The Genome Analysis Toolkit: a MapReduce framework for analyzing next-generation DNA sequencing data. *Genome Res*. Sep 2010;20(9):1297-303. doi:10.1101/gr.107524.110

5.	McLaren W, Gil L, Hunt SE, et al. The Ensembl Variant Effect Predictor. *Genome Biol*. Jun 6 2016;17(1):122. doi:10.1186/s13059-016-0974-4

6.	Wang K, Li M, Hakonarson H. ANNOVAR: functional annotation of genetic variants from high-throughput sequencing data. *Nucleic Acids Res*. Sep 2010;38(16):e164. doi:10.1093/nar/gkq603

7.	Liu X, Jian X, Boerwinkle E. dbNSFP v2.0: a database of human non-synonymous SNVs and their functional predictions and annotations. *Hum Mutat*. Sep 2013;34(9):E2393-402. doi:10.1002/humu.22376

8.	Phan L, Zhang H, Wang Q, et al. The evolution of dbSNP: 25 years of impact in genomic research. *Nucleic Acids Res*. Jan 6 2025;53(D1):D925-d931. doi:10.1093/nar/gkae977

9.	Ioannidis NM, Rothstein JH, Pejaver V, et al. REVEL: An Ensemble Method for Predicting the Pathogenicity of Rare Missense Variants. *Am J Hum Genet*. Oct 6 2016;99(4):877-885. doi:10.1016/j.ajhg.2016.08.016

10.	Alirezaie N, Kernohan KD, Hartley T, Majewski J, Hocking TD. ClinPred: Prediction Tool to Identify Disease-Relevant Nonsynonymous Single-Nucleotide Variants. *Am J Hum Genet*. Oct 4 2018;103(4):474-483. doi:10.1016/j.ajhg.2018.08.005

11.	Jagadeesh KA, Paggi JM, Ye JS, et al. S-CAP extends pathogenicity prediction to genetic variants that affect RNA splicing. *Nat Genet*. Apr 2019;51(4):755-763. doi:10.1038/s41588-019-0348-4

12.	Jaganathan K, Kyriazopoulou Panagiotopoulou S, McRae JF, et al. Predicting Splicing from Primary Sequence with Deep Learning. *Cell*. Jan 24 2019;176(3):535-548.e24. doi:10.1016/j.cell.2018.12.015

13.	Fromer M, Purcell SM. Using XHMM Software to Detect Copy Number Variation in Whole-Exome Sequencing Data. *Curr Protoc Hum Genet*. Apr 24 2014;81:7.23.1-7.23.21. doi:10.1002/0471142905.hg0723s81

14.	Talevich E, Shain AH, Botton T, Bastian BC. CNVkit: Genome-Wide Copy Number Detection and Visualization from Targeted DNA Sequencing. *PLoS Comput Biol*. Apr 2016;12(4):e1004873. doi:10.1371/journal.pcbi.1004873

15.	Fowler A, Mahamdallie S, Ruark E, et al. Accurate clinical detection of exon copy number variants in a targeted NGS panel using DECoN. *Wellcome Open Res*. Nov 25 2016;1:20. doi:10.12688/wellcomeopenres.10069.1

16.	Magi A, Tattini L, Palombo F, et al. H3M2: detection of runs of homozygosity from whole-exome sequencing data. *Bioinformatics*. Oct 15 2014;30(20):2852-9. doi:10.1093/bioinformatics/btu401

17.	Quinodoz M, Peter VG, Bedoni N, et al. AutoMap is a high performance homozygosity mapping tool using next-generation sequencing data. *Nat Commun*. Jan 22 2021;12(1):518. doi:10.1038/s41467-020-20584-4

18.	Yauy K, de Leeuw N, Yntema HG, Pfundt R, Gilissen C. Accurate detection of clinically relevant uniparental disomy from exome sequencing data. *Genet Med*. Apr 2020;22(4):803-808. doi:10.1038/s41436-019-0704-x

19.	Dolzhenko E, Deshpande V, Schlesinger F, et al. ExpansionHunter: a sequence-graph-based tool to analyze variation in short tandem repeat regions. *Bioinformatics*. Nov 1 2019;35(22):4754-4756. doi:10.1093/bioinformatics/btz431

20.	Kim YG, Lee NY, Ham JY, Lee T, Ki CS, Song KE. The Diagnostic Yield and Difficulties of Utilizing Soft-clipped Read Clusters Encountered in Clinical Exome Sequencing. *Clin Lab*. Apr 1 2023;69(4)doi:10.7754/Clin.Lab.2022.220731

21.	Ghosh R, Harrison SM, Rehm HL, Plon SE, Biesecker LG. Updated recommendation for the benign stand-alone ACMG/AMP criterion. *Hum Mutat*. Nov 2018;39(11):1525-1530. doi:10.1002/humu.23642

22.	Richards S, Aziz N, Bale S, et al. Standards and guidelines for the interpretation of sequence variants: a joint consensus recommendation of the American College of Medical Genetics and Genomics and the Association for Molecular Pathology. *Genet Med*. May 2015;17(5):405-24. doi:10.1038/gim.2015.30

23.	Kelly MA, Caleshu C, Morales A, et al. Adaptation and validation of the ACMG/AMP variant classification framework for MYH7-associated inherited cardiomyopathies: recommendations by ClinGen's Inherited Cardiomyopathy Expert Panel. *Genet Med*. Mar 2018;20(3):351-359. doi:10.1038/gim.2017.218

24.	Gelb BD, Cavé H, Dillon MW, et al. ClinGen's RASopathy Expert Panel consensus methods for variant interpretation. *Genet Med*. Nov 2018;20(11):1334-1345. doi:10.1038/gim.2018.3

25.	Shen J, Oza AM, Del Castillo I, et al. Consensus interpretation of the p.Met34Thr and p.Val37Ile variants in GJB2 by the ClinGen Hearing Loss Expert Panel. *Genet Med*. Nov 2019;21(11):2442-2452. doi:10.1038/s41436-019-0535-9

26.	Oza AM, DiStefano MT, Hemphill SE, et al. Expert specification of the ACMG/AMP variant interpretation guidelines for genetic hearing loss. *Hum Mutat*. Nov 2018;39(11):1593-1613. doi:10.1002/humu.23630

27.	Mester JL, Ghosh R, Pesaran T, et al. Gene-specific criteria for PTEN variant curation: Recommendations from the ClinGen PTEN Expert Panel. *Hum Mutat*. Nov 2018;39(11):1581-1592. doi:10.1002/humu.23636

28.	Abou Tayoun AN, Pesaran T, DiStefano MT, et al. Recommendations for interpreting the loss of function PVS1 ACMG/AMP variant criterion. *Hum Mutat*. Nov 2018;39(11):1517-1524. doi:10.1002/humu.23626

29.	Lee K, Krempely K, Roberts ME, et al. Specifications of the ACMG/AMP variant curation guidelines for the analysis of germline CDH1 sequence variants. *Hum Mutat*. Nov 2018;39(11):1553-1568. doi:10.1002/humu.23650

30.	Zastrow DB, Baudet H, Shen W, et al. Unique aspects of sequence variant interpretation for inborn errors of metabolism (IEM): The ClinGen IEM Working Group and the Phenylalanine Hydroxylase Gene. *Hum Mutat*. Nov 2018;39(11):1569-1580. doi:10.1002/humu.23649

**eFigure**

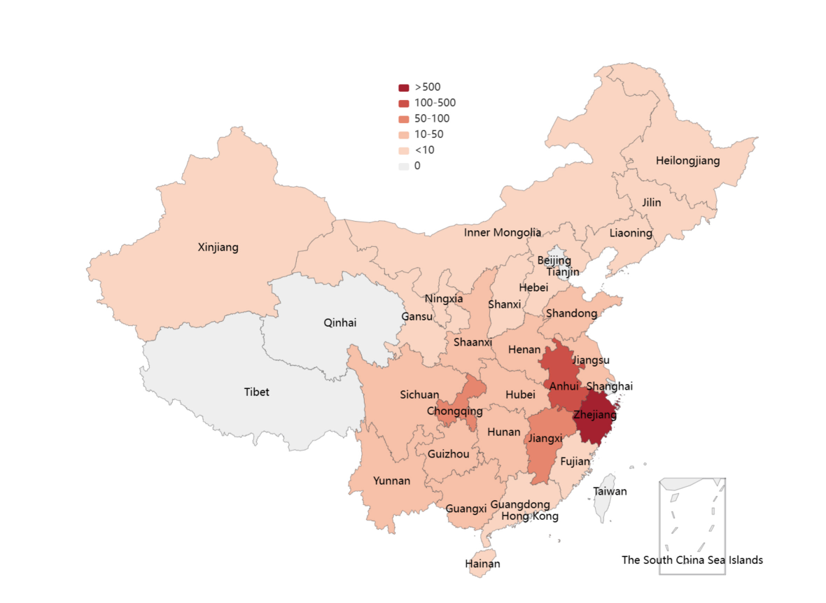

**eFigure 1**. **The geographical distribution of participants in this study.** According to their ID registration information, the geographic distribution of 1382 singleton pregnant women met the inclusion criteria and consented to participate in this study was illustrated. Though the majority of study subjects were from Zhejiang, Anhui, Jiangxi and Chongqing, the participants still represented diverse social-economic developments in this study.

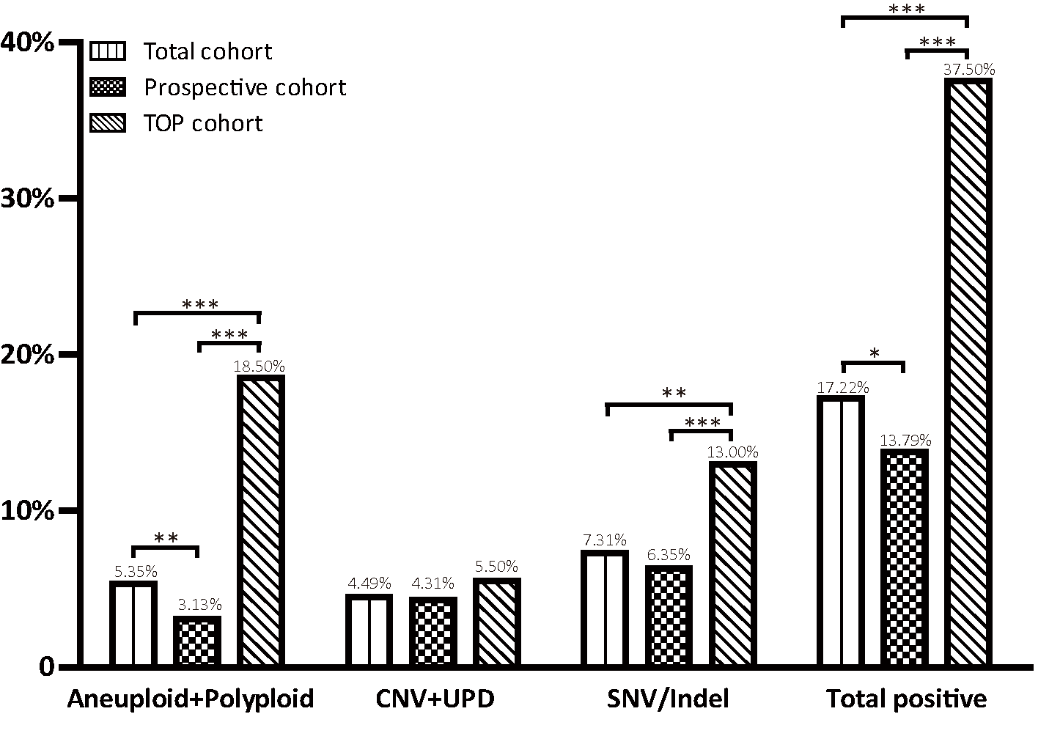

**eFigure 2.** **Comparison on the positive rate of different types of variations in different cohorts.** The total cohort consisted of 1382 singleton pregnancies from seven prenatal diagnosis centers across China from October 2022 to April 2024 strictly according to the inclusion criteria of this study. Amongst, 200 pregnancies terminated prior to genetic testing were designated as the TOP cohort. The remaining 1182 cases was taken as the prospective cohort. The numbers displayed on top of each bar represent the positive rate of a specific variant, while the labels along the x-axis detail the types of variations and the total variants. CNV: copy number variation; UPD: uniparental disomy; SNV: single nucleotide variant; Indel: insertion/deletion. *p＜0.05；** p＜0.01; *** p＜0.001

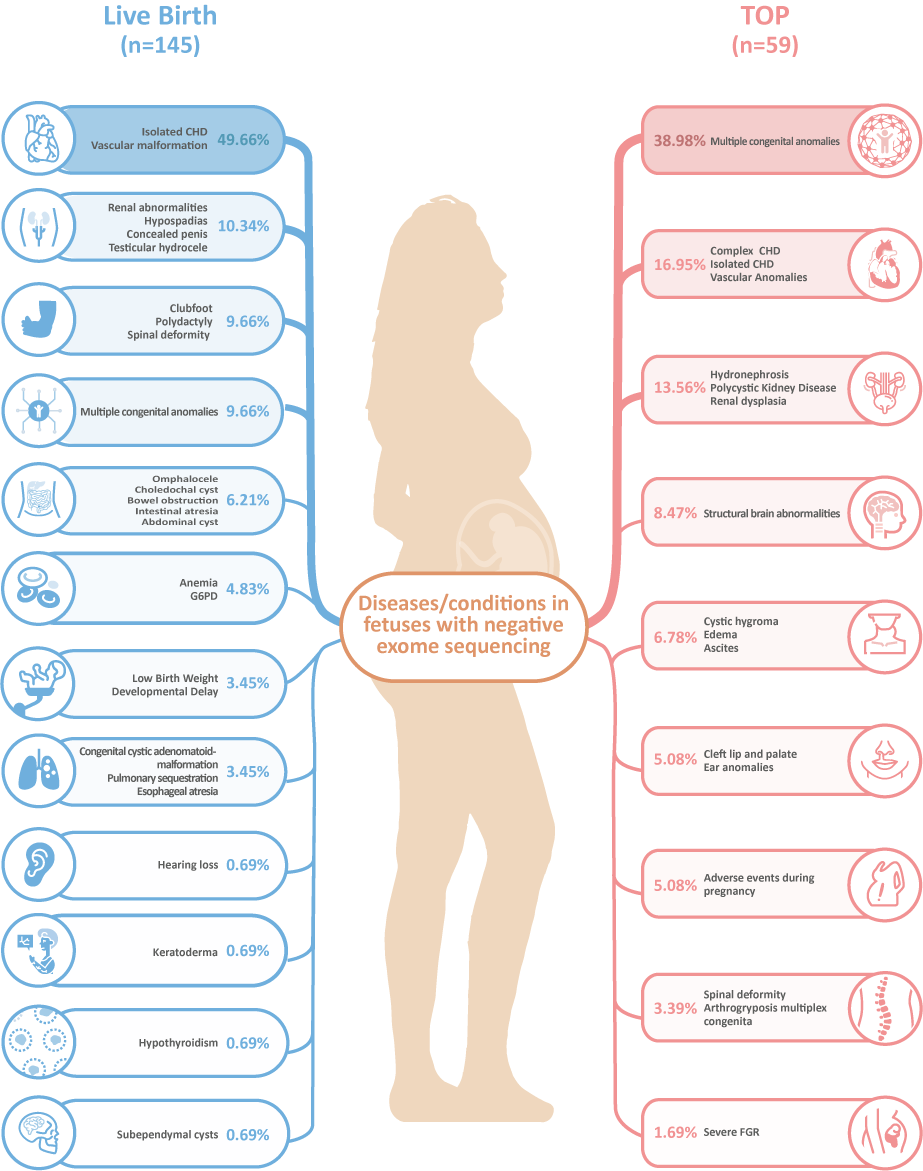

**eFigure 3.** **Comparison on spectrum of diseases/conditions between fetuses who were born alive and those with TOP in negative trio-WES population.** It’s revealed that the decision-makings for pregnancies who were tested negative in trio-WES was significantly influenced by the disease severity or clinical actionability, in addition to genetic testing results. For instance, 72 of 145 (49.66%) fetuses with isolated congenital heart disease (CHD) were born alive, while 23 pregnancies (38.98%) with fetal multiple congenital anomaly (MCA) and 10 pregnancies (16.95%) with fetal complex CHD were terminated.
